# Individual and spatial determinants of mortality during the Covid-19 pandemic: The case of Belgium in 2020

**DOI:** 10.1101/2023.09.04.23295014

**Authors:** Mélanie Bourguignon, Aurélie Bertrand, Joan Damiens, Yoann Doignon, Thierry Eggerickx, Audrey Plavsic, Jean-Paul Sanderson

## Abstract

**Context:** The year 2020 was marked by the Covid-19 pandemic. In Belgium, it led to a doubling in deaths, mainly grouped into two periods. This article aims to compare the relative importance of predictors and individual and spatial determinants of mortality during these two waves to an equivalent non-pandemic period and to identify whether and to what extent the pandemic has altered the sociodemographic patterns of conventional mortality.

**Methods:** The analyses relate to all-cause mortality during the two waves of Covid-19 and their equivalent in 2019. They are based on matching individual and exhaustive data from the Belgian National Register with tax and population census data. A multi-level approach was adopted combining individual and spatial determinants.

**Results:** Mortality patterns during the pandemic are very similar to those observed outside the pandemic. As in 2019, age, sex, and household composition significantly determine the individual risk of dying, with a higher risk of death among the oldest people, men, and residents of collective households. However, their risk of death increases during the Covid period, especially in the 65–79 age group. Spatial information is no more significant in 2020 than in 2019. However, a higher risk of death is observed when the local excess mortality index or the proportions of isolated or disadvantaged people increase.

**Conclusions:** While the Covid pandemic did not fundamentally alter conventional mortality patterns, it did amplify some of the pre-existing differences in mortality.

## Introduction

The Covid-19 pandemic was the deadliest episode in the history of Belgium since the Second World War, incurring more than 33,000 deaths between 2020 and 2022. Regardless of the method used to count deaths^1^, which varies from one country to another, Belgium appears to be one of the countries where the mortality rate from Covid-19 in 2020 was particularly high (Bustos Sierra et al., 2020; Sánchez-Páez, 2022). Two main waves of excess mortality have been identified (Figure 1). The first ran from mid-March to the end of April 2020 and the second from mid-October to the end of December 2020. Both were characterised by a doubling in the number of deaths compared with a so-called ‘normal’ situation (the monthly average of deaths observed between 2016 and 2019). This translates into a loss of one year of life expectancy at birth in Belgium between 2019 and 2020 (Bourguignon et al., 2022), one of the most significant international losses (Aburto et al., 2021; Islam et al., 2021).

**Figure 1.**
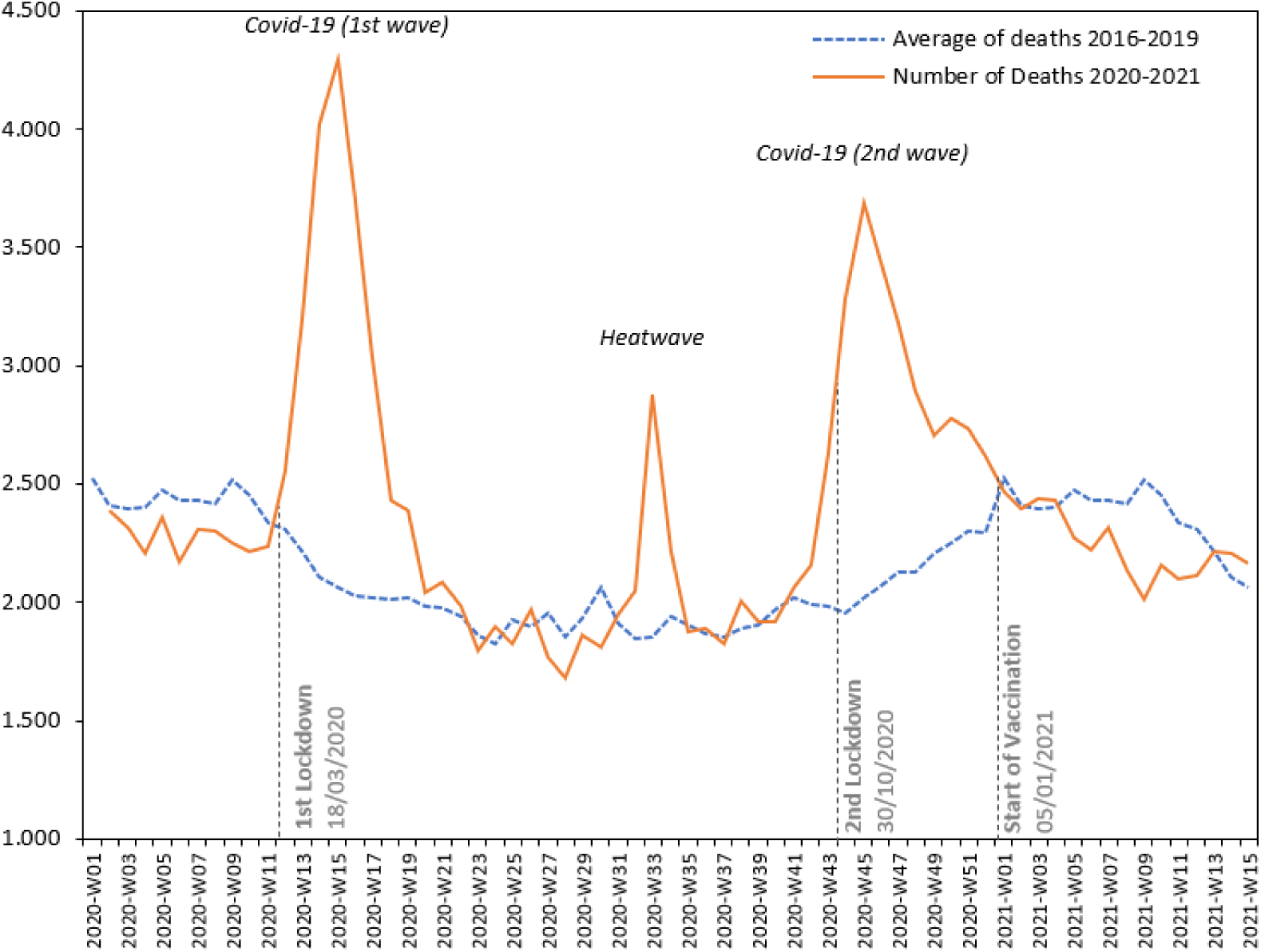
Trends in excess mortality linked to Covid-19 in Belgium. Source: Statbel, authors’ calculations.

In Belgium and worldwide, very few studies have looked at the spatial determinants of mortality during a pandemic. Moreover, their relative importance compared with individual determinants remains largely unknown. Is Covid-19-related mortality unique, or does it follow the usual trends? ‘Spatial determinants’ and ‘contextual determinants’ are used synonymously and refer to the characteristics of the environment in which individuals live (see section ‘Hypotheses, data and methods’).

### Individual determinants

Studies agree on the major role of age in the mortality caused by the pandemic: as age increases, so does the risk of mortality from Covid-19, particularly in high-income countries (Bauer et al., 2021; Guilmoto, 2020; Islam et al., 2021; Pifarré i Arolas et al., 2021; Williamson et al., 2020). In Belgium, the increase in excess mortality with age is highly conspicuous, particularly during the first wave of the pandemic, especially from the age of 65 onwards (Bourguignon et al., 2022). This can be explained in particular by health status: as age increases, health status deteriorates (Bambra et al., 2020), and the risk of contamination and death linked to Covid-19 is greater in people with co-morbidities such as cardiovascular disease, diabetes, cancer, and lung disease (Atkins et al., 2020; Burström & Tao, 2020). During the second wave, the gradient of excess mortality by age persists but is less clear-cut, with the situation for those over 85 years of age approaching that of the 75–84 age group (Bourguignon et al., 2022).

The high excess mortality among the elderly in 2020 also depends on where they live. In many countries, residential institutions for the elderly were places where Covid-19 mortality was particularly high (Brandén et al., 2020; Kemenesi et al., 2020). In Belgium, during the first wave, they accounted for 50% of deaths linked to Covid-19 related deaths (Sciensano, 2020). In Wallonia (the southern region of Belgium), 65% of deaths involved individuals staying in these institutions, whereas only 1.3% of the Walloon population resides in such establishments (Hardy et al., 2021). For the country as a whole, the standardised mortality rate for people living in collective households in 2020 was 1.7 times higher than the same rate calculated for the 2015–2019 period in the first wave and 1.5 times higher in the second wave (Bourguignon et al., 2022). This excess mortality is due to the age and state of health of the residents but also to the proximity in these residences between individuals and with carers (Davidson & Szanton, 2020; Kemenesi et al., 2020). The health crisis highlighted structural weaknesses, including a lack of resources (e.g., face masks) and staff that these establishments were experiencing.

Even within the home, household composition may have had an impact on the risk of dying from Covid-19, given the importance of interpersonal contact in transmitting the virus. According to a study of people over 65 living at home in England (Nafilyan et al., 2021), living in a multigenerational household is associated with a higher risk of death (compared with a two-adult household). Another study in Los Angeles in the United States showed a positive association between household size and the risk of dying from Covid-19 (Varshney et al., 2022). In the case of Stockholm, the risk of dying from Covid-19 was higher for elderly people living with adults in working age than for those living alone (Brandén et al., 2020).

Furthermore, the risk of dying from Covid-19 was higher in men than in women at any age in most countries (Aburto et al., 2021; Ahrenfeldt et al., 2021), including Belgium (Gadeyne et al., 2021). In general, the gender gap increases until the 60–69 age bracket, then gradually decreases to become very small after the age of 80 (Ahrenfeldt et al., 2021). This could be explained by a selection effect linked to the health of men and women at different ages.

The influence of nationality on the risk of death from Covid-19 is less definite. Some studies have shown that people of foreign nationality were affected by higher excess mortality during the pandemic period than natives, as was the case in Sweden, irrespective of age and socio-economic characteristics (Drefahl et al., 2020), and in the United States and the United Kingdom (Abedi et al., 2021; Andrasfay & Goldman, 2021; Williamson et al., 2020). In Italy, on the other hand, mortality patterns for migrants and non-migrants in 2020 did not differ significantly from those observed in the preceding years (Canevelli et al., 2020).

Over and above age, gender, household type, and nationality, several studies have shown that Covid-19 hit the disadvantaged social classes harder (Abedi et al., 2021; Andrasfay & Goldman, 2021; Barhoumi et al., 2020; Chen & Krieger, 2021; Mayer, 2022; Williamson et al., 2020). Firstly, this may be due to the increased presence of co-morbidities at an earlier age among the most disadvantaged populations, which already increases the risk of dying from Covid-19 (Bambra et al., 2020; Williamson et al., 2020). Disadvantaged populations are characterised by more widespread risk behaviours (smoking, poor dietary habits), less understanding of and adherence to health and hygiene measures to combat the disease, and limited access to and/or use of good quality healthcare. (Gadeyne et al., 2021). Secondly, socially disadvantaged populations are more frequently found in densely populated neighbourhoods and densely occupied housing, which are additional factors of exposure to the risk of infection and, by extension, mortality (Bambra et al., 2020; Barhoumi et al., 2020). Thirdly, disadvantaged populations are over-represented among ‘front-line’ occupations, which have not benefited as much from the protective effects of containment and teleworking (Bambra et al., 2020; Barhoumi et al., 2020).

In Belgium, the few studies considering the social dimension of Covid-19 present variable results depending on the methodologies and social-positioning indicators used. Decoster et al. (2021) showed the existence of a negative income gradient in terms of excess mortality among the elderly in Belgium in the first wave (Decoster et al., 2021). Conversely, Gadeyne et al. (2021), using income and level of education as indicators of social inequality, identified the highest levels of excess mortality among the highest-income men aged 25–64 and among middle- and high-income men and women aged 65–84. Bourguignon et al. (2022) found excess mortality among all social groups^2^ based on an excess mortality index^3^ calculated for the two Covid-19 waves in 2020. However, the most disadvantaged social group shows the highest level of excess mortality, with a visible social gradient that is more marked for the population aged 40–79 than for those aged 80 and over, especially in the second wave. The significant fall in life expectancy in 2020 is more marked for the most disadvantaged social group (Bourguignon et al., 2022).

### Spatial and contextual determinants

Like other epidemics, Covid-19 has a particular spatial structure (Amdaoud et al., 2021). In addition to individual determinants, certain characteristics of the place of residence can impact the individual risk of dying from Covid-19. Population density and mobility play a major role in the spread of the virus (Fonseca-Rodríguez et al., 2021). In the United States, among the seven states most affected, the most densely populated counties were the ones with the highest rates of contamination (Abedi et al., 2021). In France, the departments with the highest urbanisation rates experienced the highest excess mortality during the first wave of Covid-19 (Pilkington et al., 2021). The contagious nature of Covid-19 means that the closer an area is to a source of contamination, the greater the probability that it will be affected by the epidemic (Amdaoud et al., 2021). Spread by contagion has been observed in France’s departments (Levratto et al., 2020) and in China’s provinces (Kang et al., 2020). Containment has proved to be an effective solution to limit this contagion, as demonstrated in France (Gaudart et al., 2021). In New York and Chicago, the ‘cold spots’ identified based on diagnosed cases were distinguished by their ability to maintain physical distancing (Maroko et al., 2020).

At the same time, there are inequalities between regions in terms of access to healthcare. If an area has a smaller and more dispersed healthcare system, this implies a lower or slower response capacity in critical situations. At the French departmental level, a strong association was found between high levels of excess mortality and poor primary care provision during the first wave of the epidemic (Pilkington et al., 2021). The social and residential environment also has an influence on exposure to the virus and the probability of dying. High population densities (Andersen et al., 2021; Fonseca-Rodríguez et al., 2021) and neighbourhoods with a high proportion of overcrowded housing (Maroko et al., 2020), immigrant or ethnic minority populations (Andersen et al., 2021; Fonseca-Rodríguez et al., 2021), and disadvantaged populations (Riou et al., 2021; Vandentorren et al., 2022) are frequently cited as contextual factors in infection and excess mortality. The association between these factors makes it difficult to understand the geography of Covid-19-related mortality. In Belgium, a significant association has been detected at the municipal level between the number of *infections* and social precariousness (Meurisse et al., 2022). Still, nothing of the kind has yet been shown concerning *excess mortality* from Covid-19.

Mortality linked to Covid-19 was not uniform across Belgium. Some areas were affected earlier and/or more severely than others, revealing the presence of outbreaks, as in other countries (Figure 2). During the first wave, the main outbreaks were in the Mons region (south-west), the province of Limburg and eastern Flemish Brabant (north-east), a large part of the province of Liège and the north-east of the province of Luxembourg (centre-east) and the Brussels conurbation (centre). But the geography of the excess mortality in the first wave differs greatly from that of the second wave, with outbreaks mainly located along international borders. Furthermore, to date, no clear link has been detected between the excess mortality linked to Covid-19 and other factors such as population density, the country’s social geography or ‘normal’ mortality, particularly during the second wave (Bourguignon et al., 2021).

**Figure 2.**
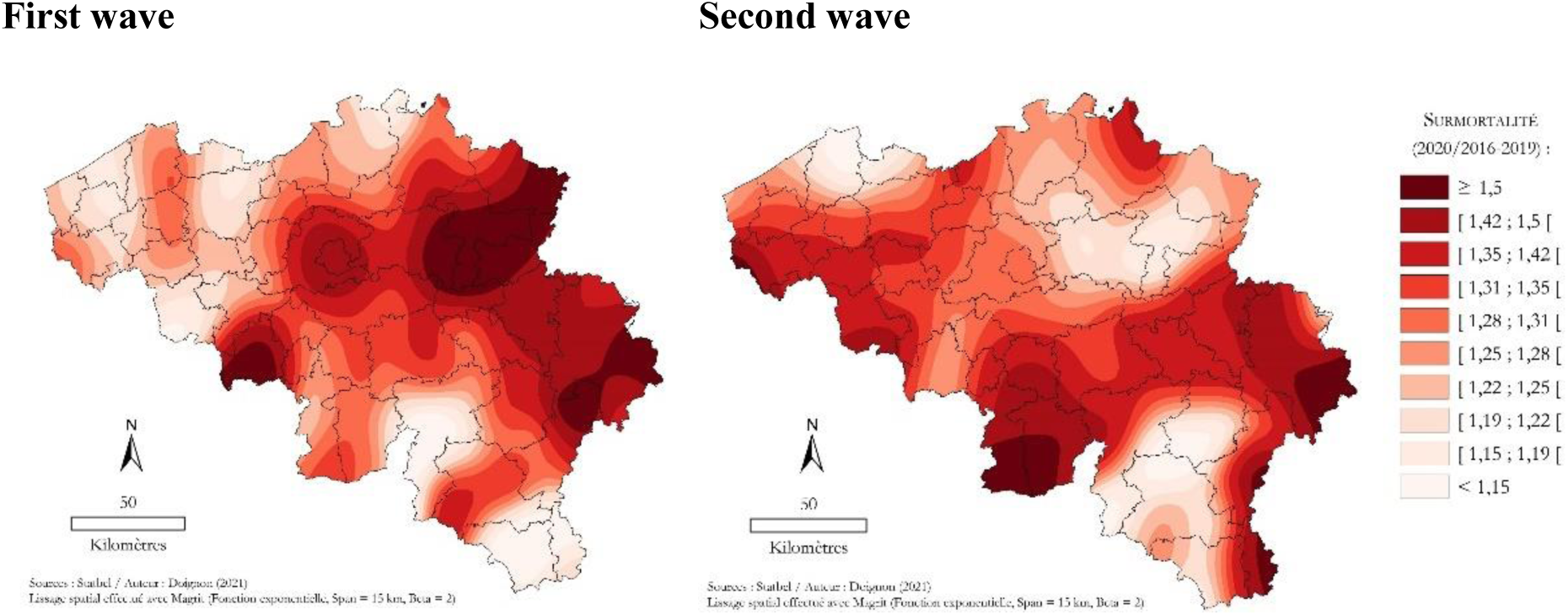
Excess mortality in the first wave (18 March–17 June 2020) and the second wave (28 September 2020–31 January 2021)^4^. Source: Statbel; authors’ calculations.

## Hypotheses, data and methods

### Research questions and hypotheses

This article has two objectives. Firstly, we measure – and are the first to do so, to our knowledge – the relative importance of the various individual and spatial determinants by ranking their weight in terms of the risk of dying during the two Covid-19 waves in 2020. Are the factors most correlated with mortality during the Covid-19 period the same as those outside the period? Secondly, we measure the differences in mortality between the different population groups defined by these determinants during the periods of excess mortality linked to Covid-19. Which characteristics are the most associated with excess mortality during the pandemic period?

These two objectives are part of a comparative perspective. Firstly, the analyses compare two types of mortality patterns, ‘classic’ (2019) and ‘pandemic’ (2020), aiming to identify potential differences between the two. The analyses are carried out separately for each Covid wave in 2020, systematically compared with the two equivalent periods in 2019. Finally, the analyses cover the whole population and three major age groups (40–64, 65–79, 80+), which may have been affected differently by the pandemic.

Our analyses and reflections are based on three hypotheses.

Firstly, the relative importance of individual and contextual determinants would change in a pandemic context. The **weight of contextual variables** would be greater than that observed in conventional mortality, given the contagious nature of the pandemic and exposure factors more closely linked to population density or nearby outbreaks.

Secondly, certain population groups would see their risk of death changed during a pandemic: the association between some **characteristics** and mortality would increase or decrease compared to a ‘classic’ mortality pattern. The vulnerability of people who are already fragile in a context of traditional mortality would be amplified during a pandemic, and this would particularly affect the elderly, men, disadvantaged social groups and residents of collective households. Conversely, isolated households would be less vulnerable in a pandemic due to their isolated lifestyle.

Thirdly, individual and contextual determinants during the pandemic would have a different effect depending on **the individual’s age**. Among older and very old adults, poor health status would increase the individual risk of dying during one of the 2020 Covid-19 waves. This would be reflected in a risk of death that increases with age or a higher risk of death for residents of collective households (of the MR(S)^5^ type). Among working-age adults, social class would play a decisive role, with an increased risk of death during the two 2020 pandemic waves among the most disadvantaged groups due to poorer health status, more frequent risk behaviours, less use of healthcare and socially discriminatory measures during periods of confinement^6^.

### Data

Several types of data were used in this article. As the analysis was conducted at the individual level, we used an anonymised individual database (Demobel) that exhaustively covers Belgium’s entire legally domiciled population, i.e., over 11 million individuals. This database, developed by the Belgian statistical office (Statbel), is the result of linking the individual-level censuses (2001 and 2011), the National Register (in which every legal resident in Belgium is entered), and the tax register for the period 1992–2022. The censuses and the tax register provide information on the socio-economic characteristics of individuals (education, occupation, housing conditions, tax income). The National Register provides information on their socio-demographic characteristics (age, gender, type of household, place of residence, etc.). This database can also identify people who died during the period covered by the data, with the date of death indicated where appropriate. This information makes it possible to identify individuals who died during the two periods of Covid-related excess mortality covered by our article.

Our approach is based on an analysis of all-cause mortality. Our analyses focus on the months marked by excess mortality associated with Covid-19. The first wave of deaths associated with the pandemic runs from March to May 2020 and the second wave from October to December 2020. Particular care has been taken to define the population at risk and its characteristics (household composition, commune of residence) at the time of each wave. For the first wave, the population at risk of dying is the population that survived at least until March. It was assigned the characteristics declared on 1 January 2020. The population observed in the second wave includes residents of Belgium who were still alive in October. To be more precise, we have associated the characteristics on 1 January 2021 with survivors only to take account of changes made during 2020 regarding household composition or place of residence, for example. For people who died in 2020, we have no choice but to use the characteristics observed on 1 January 2020. The same procedure was followed to define the populations at risk of dying in 2019.

### Methods

Our questions and hypotheses were tested using a two-pronged approach: firstly, to identify the individual and contextual determinants of mortality during the Covid-19 pandemic, and secondly, to measure the relative importance of these determinants. To do this, we model the individual probability of dying during each of the two waves^7^ of excess mortality during Covid-19. As we wish to distinguish the effect of individual variables from that of contextual variables, we estimate a multilevel random-effects logistic model:

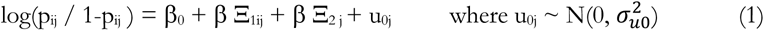

where p_ij_ denotes the probability of an individual *i* residing in a commune *j* dying during a Covid-19 wave; X_ij_ a vector of individual explanatory variables; X_j_ a vector of contextual explanatory variables; u_0j_ a random residual term at the level of the groups defined by the communes (this term groups together the unobservable random effects at the commune level).

Several variables are included in the multilevel models to explain the probability of dying during a wave of excess Covid-19 mortality. These variables are either individual or contextual. **Individual variables** include (1) the age of individuals, (2) their sex, (3) the composition of their household, (4) their nationality, and (5) their social background.

A multidimensional indicator of social precariousness captures the socioeconomic category of individuals based on four dimensions that traditionally enable individuals to be positioned socially (Cambois & Jusot, 2007; Kunst & Mackenbach, 1994): education, employment category, and housing conditions from the 2011 census, and household income from the 2017 tax returns. While the correlation between these dimensions is high, they each play a different role in health status and mortality. Level of education measures cultural capital. It will determine attitudes toward preventative behaviours, use of healthcare, and access to healthcare. Income, employment category, and housing conditions refer more to material resources and standard of living (Cambois & Jusot, 2007). This multidimensional index considers each individual’s overall situation and has been divided into score quartiles: disadvantaged, low intermediate, high intermediate, and advantaged (Eggerickx et al., 2020). A fifth group was observed: the ‘undetermined’, for whom no information is available for at least two of the dimensions considered, either because they did not answer the questions or were not present at the time of the census (e.g. children born or immigrants arrived in after 2011).

At the same time, **contextual variables** relating to the characteristics of individuals’ place of residence are included: (1) the proportion of disadvantaged people in the commune of residence, (2) the commune’s population density, (3) the proportion of isolated people, (4) the level of excess mortality in all communes within 20 kilometres of the commune of residence (incl.), calculated by comparing the mortality observed in the two 2020 waves with the average for the years 2016–2019 at the same time, (5) the distance to the nearest hospital^8^, and (6) the position of the commune of residence in the country’s urban hierarchy^9^.

In total, 16 models are produced for the total population and three main age groups (40–64, 65– 79, 80+)^10^. The use of these population subgroups will enable us to compare the relationships of each predictor with the risk of death, and to see whether or not the directions and intensity of these relationships are identical. Nevertheless, the comparison of estimates must be made with caution, due to the existence of unobserved heterogeneity: the variation in mortality from one age group to another may depend on unobserved variables, and this unobservable variation may be different (Mood, 2010). Each of the four main models is reproduced for each of the two waves in 2020 and for the two equivalent periods in 2019 (base year) (Figure 3)

**Figure 3:**
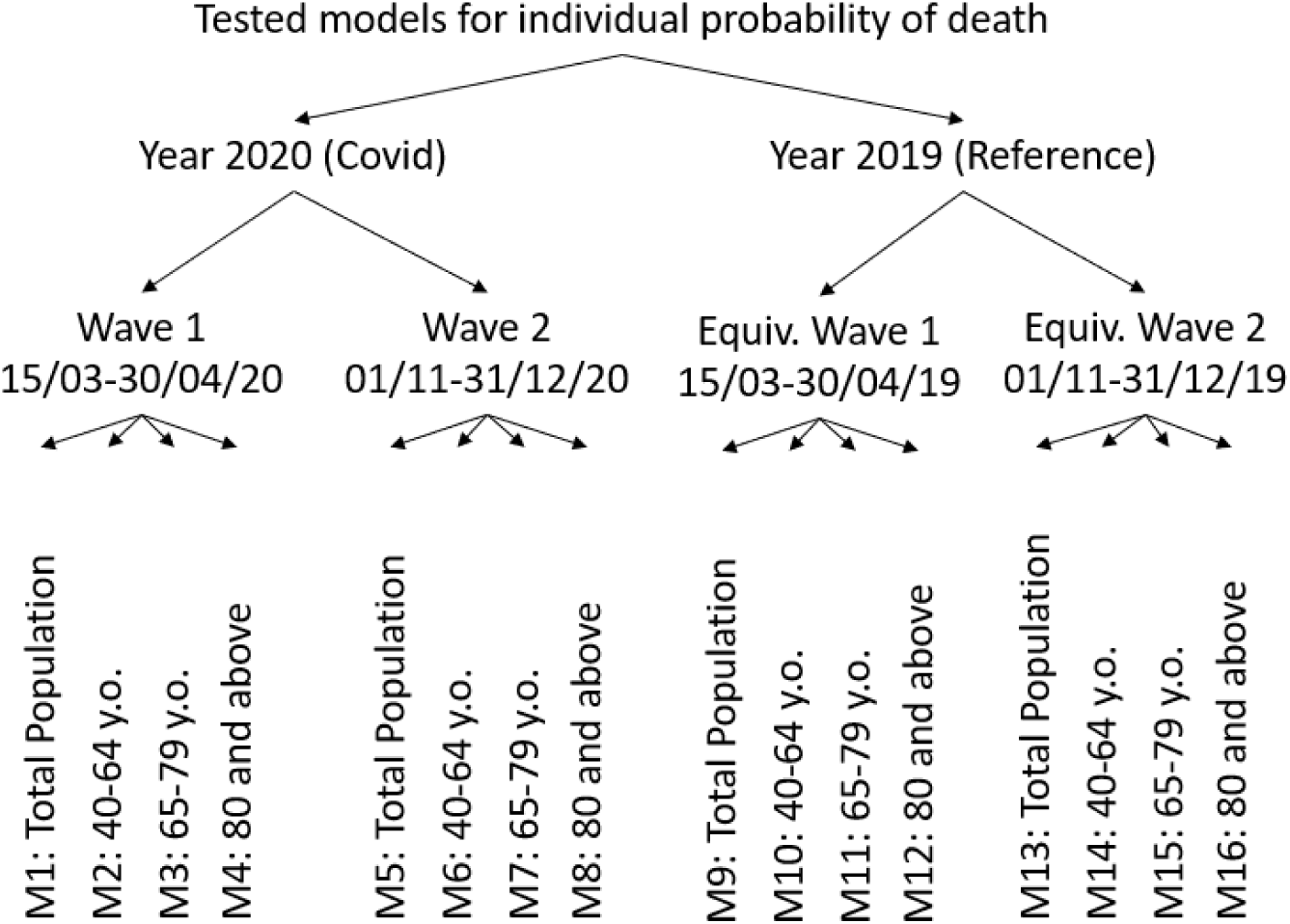
Estimated multilevel models

Analysis of the regression models makes it possible to identify the individual and contextual determinants of the individual risk of death. Measuring the relative importance of the different explanatory variables involves analysing the marginal contributions of the predictors in reducing the unexplained variability of mortality, all other things being equal (for a fixed value of the other variables introduced into the model). For each explanatory variable, we calculated a partial pseudo-correlation (*R*) adapted to the logistic model (Bhatti et al., 2006):

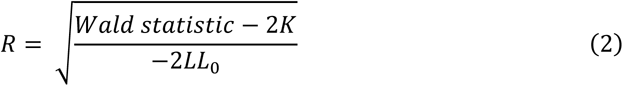

Where *K* is the number of degrees of freedom of the explanatory variable and *LL* is the log-likelihood of the empty model without independent variables. Partial pseudo-correlation is interpreted similarly as a correlation coefficient (between −1 and 1). In our case, the absolute value of the indicator will be used, as we are interested in the intensity of partial pseudo-correlations independently of their direction.

### Analysis of an exhaustive population

The data we used were drawn from the National Register and population censuses. The population studied is not a representative sample of Belgium’s resident population but rather is the total population residing in Belgium and included in the National Register. Registration in these databases is compulsory, and in theory, only illegal migrants, asylum seekers, and diplomatic personnel are not covered by this data source. Given the exhaustiveness of the data, this research does not fall within the inferential statistics framework. We do not consider that there is a ‘super-population’ from which our study population is drawn or that the latter is one possible reality among others. By definition, inferential statistical tools should be used when the data constitute a sample and the results are expected to be generalised to a larger population (White & Gorard, 2017), which is not the case here.

Thus, the *b* parameters derived from the regression models are not an estimate of the sample parameter to which a confidence level is assigned. These parameters are real and represent the effect of an explanatory variable on the variable of interest in the population studied. In this way, we do not interpret using the tools of inferential statistics (*p*-value, confidence intervals). Instead, we systematically use the number of individuals and the proportion of deaths for each modality of each categorical variable used. This sometimes means we can temper our comments, for example, if the analyses involve too small a group of individuals.

## Analysis

### Relative importance of variables

In 2020, during the two waves of the Covid-19 pandemic, the individual probability of dying was mainly influenced by age, household type, sex, and local excess mortality. In comparison, the weight of contextual variables is very low overall, except for excess local mortality during the pandemic period. Age is the factor with the greatest relative importance for the probability of dying during the two Covid waves in 2020. In the first wave, it was 2.5 times more important than the household composition variable and 5 times more important than the sex variable (Figure 4). This finding is unsurprising and is also observed for mortality outside any pandemic episode. More surprising is the low weight of certain individual variables, such as social group and nationality, both during and outside the pandemic.

**Figure 4:**
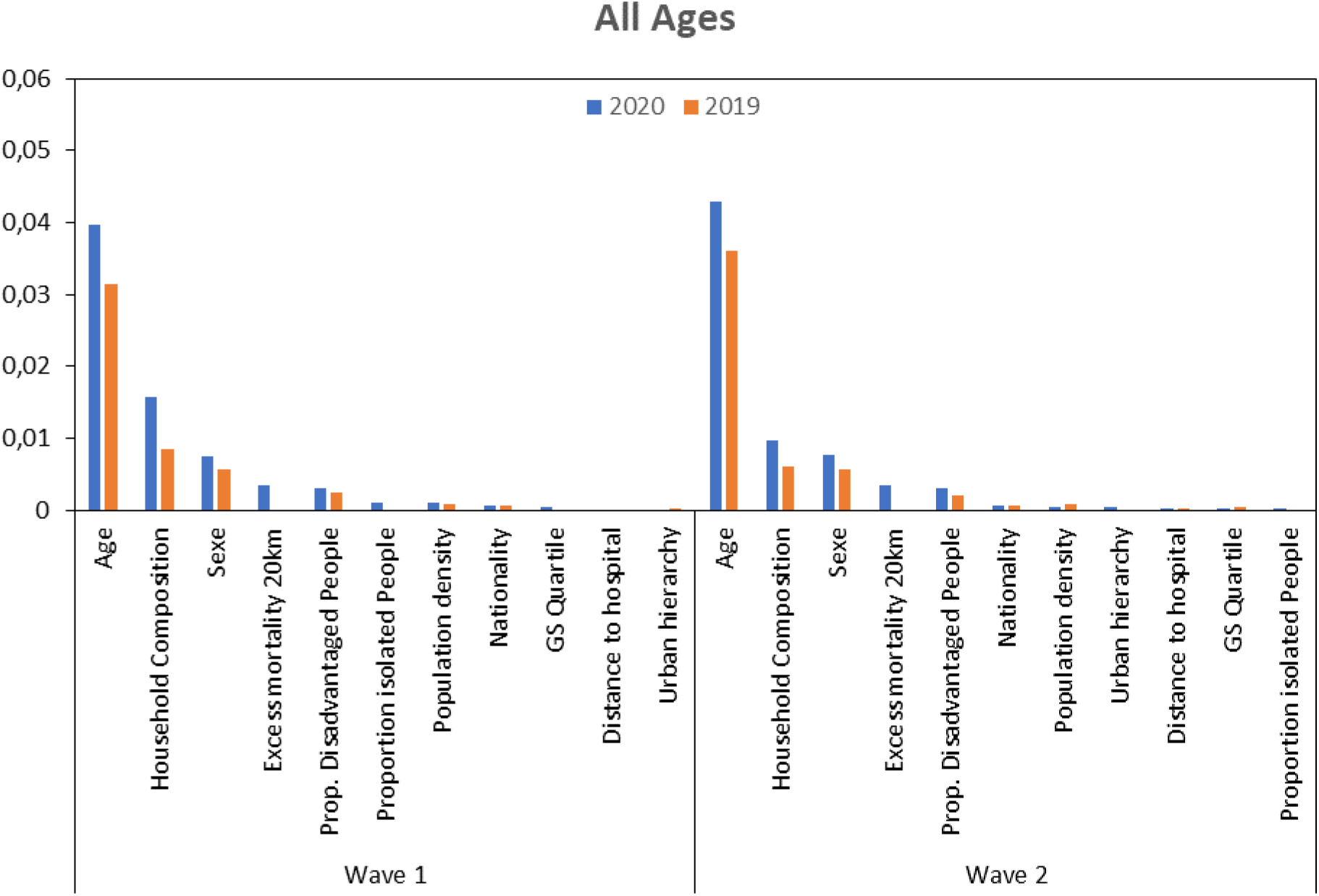
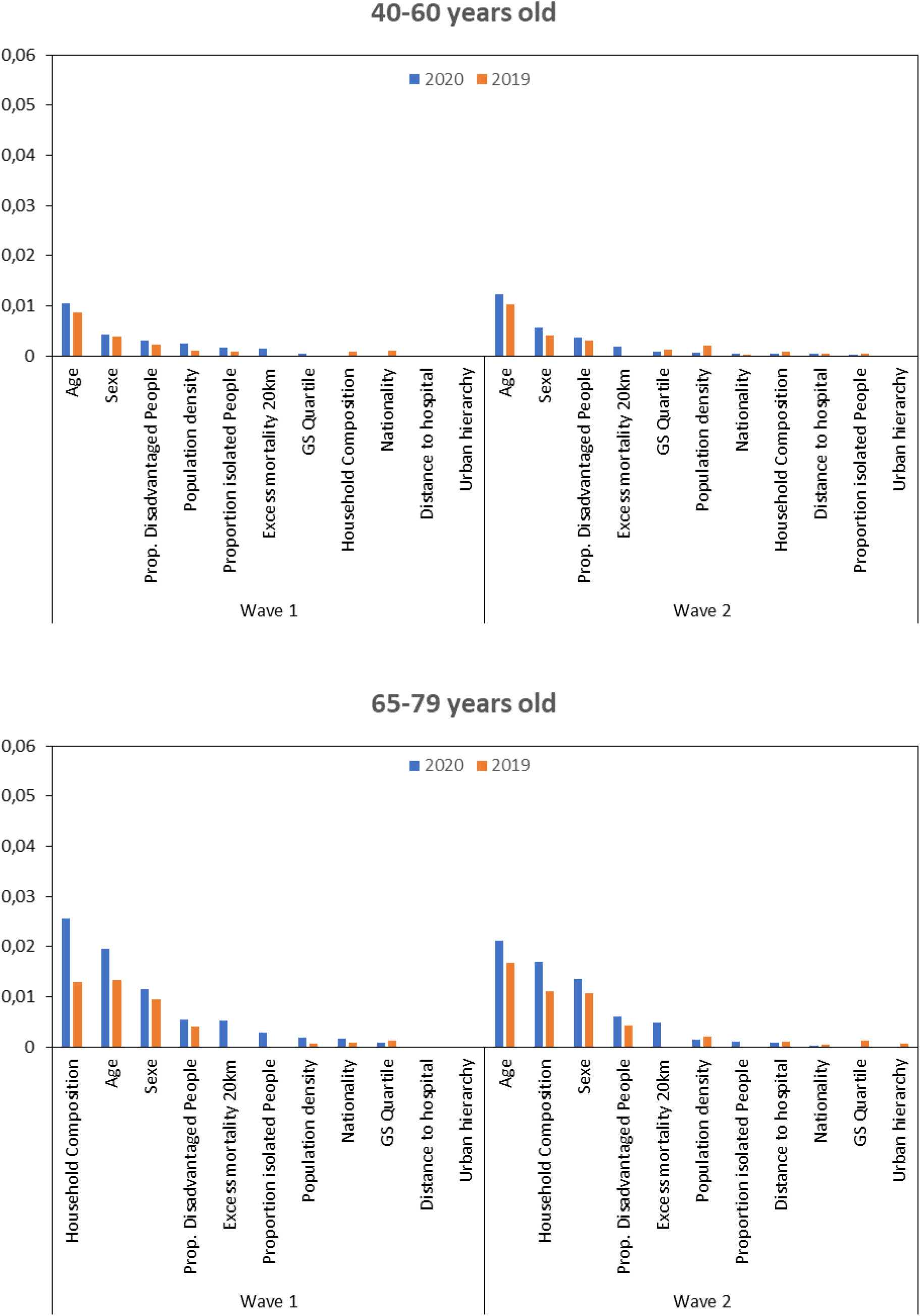
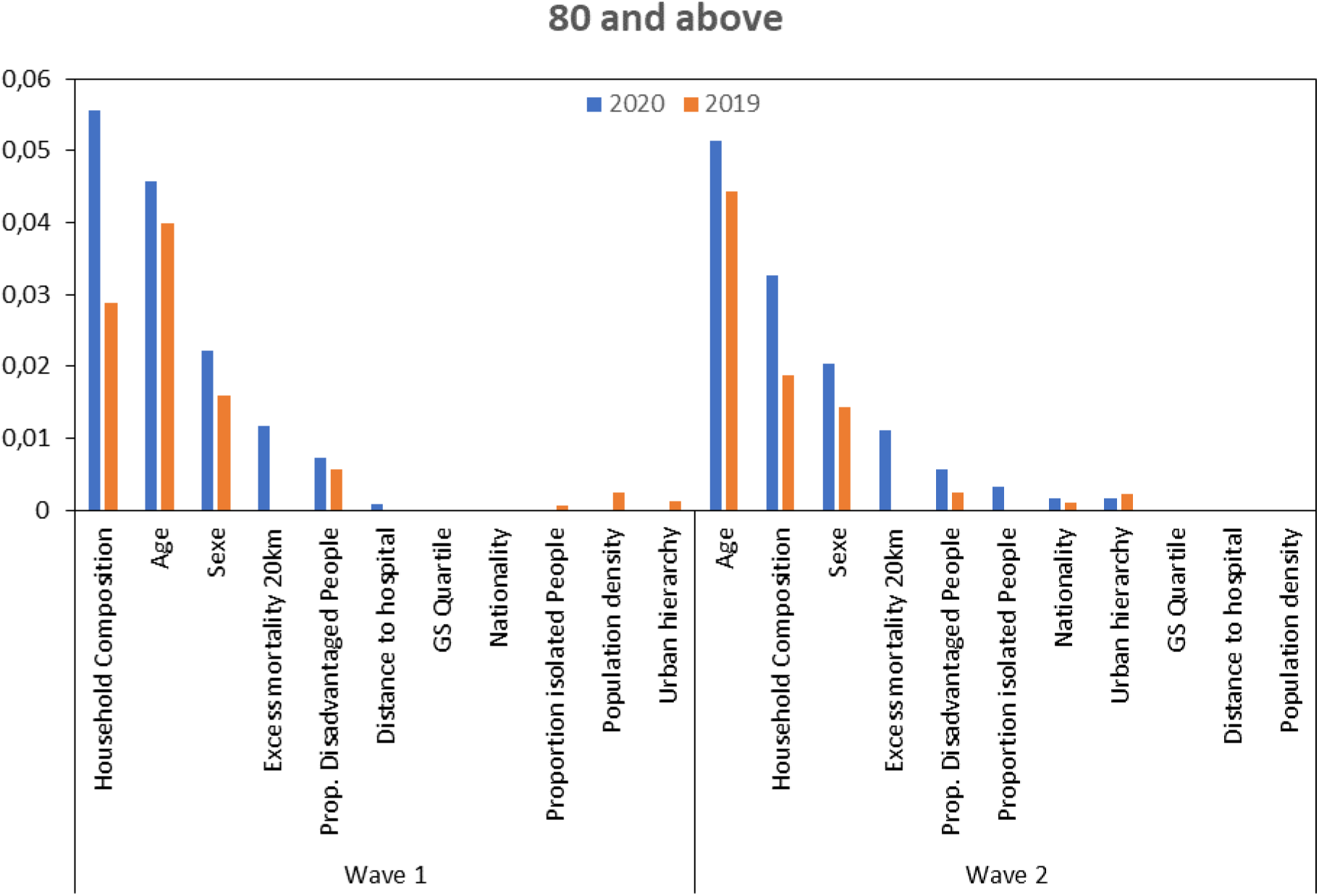
Partial pseudo-correlations from multilevel models estimating the probability of dying during the first or second wave (2020) of the Covid-19 pandemic and the equivalent of these periods for 2019. Graphs produced based on ‘all ages’ models and by major age groups; prioritisation of variables according to their respective weight in the models for the year 2020. Source: Statbel; authors’ calculations.

### Relative importance of variables by age group

There are nonetheless slight differences between the main age groups. Among 40–64-year-olds, age and gender retain their high relative weight, while that of household composition declines. Conversely, contextual determinants – the proportion of disadvantaged or isolated households in the municipality, population density, and local excess mortality – are better positioned in the ranking than for older age groups, even if their relative weight remains low. For older people (aged 65–79 or 80 and over), the variables that have a decisive influence on individual mortality are relatively different. During the first wave of the Covid-19 pandemic, household composition had the highest weight, followed closely by age and sex. The dominance of household composition underlines, in particular, the devastating effect of the epidemic in nursing homes. In contrast, for ‘usual’ mortality (2019), the decisive role is attributed to the age of individuals. In the second wave, the situation ‘normalises’, with the return of age as the leading variable in terms of partial pseudo-correlations.

Finally, it should be noted that, except for the 40–64 age group in the first wave, excess local mortality was among the five variables with the highest relative importance. This means that in the context of age-related frailty (and therefore health-related frailty), excess mortality in the surrounding area is a determining (or aggravating) factor in mortality during the Covid period.

### Characteristics related to mortality

The all-ages models (Table 1) show that men, older people, disadvantaged social categories, and certain types of households, such as collective households and single people, have a higher mortality rate than other categories, both during the Covid period and outside the pandemic. In 2020, in both waves, women were half as likely as men to die (in wave 1, Odds-Ratio (OR) = 0.523, and in wave 2, OR = 0.553). This factor is just as strong outside the pandemic (in wave 1, OR 2019 = 0.547, and in wave 2, OR = 0.592). Men’s vulnerability was therefore not a distinctive feature of mortality during Covid-19.

**Table 1.** Multivariate results estimating the individual probability of dying (expressed in odds ratios) during one of the two Covid-19 waves (2020), compared with the results obtained during the same periods in 2019, models all ages combined. Source: Statbel; authors’ calculations.

|  | MODEL 1: All – V1 2020 |  |  | MODEL 9: All – V1 2019 |  |  | MODEL 5: All – V2 2020 |  |  | MODEL 13: All – V2 2019 |  |  |
| --- | --- | --- | --- | --- | --- | --- | --- | --- | --- | --- | --- | --- |
|  | OR | Population | Number of deaths per 100,000 | OR | Population | Number of deaths per 100,000 | OR | Population | Number of deaths per 100,000 | OR | Population | Number of deaths per 100,000 |
| **Sex (ref. Male)** |  | **5,660,064** | **187.4** |  | **5,628,228** | **122.9** |  | **5,649,456** | **225.5** |  | **5,621,309** | **160.5** |
| Female | 0.523 | 5,832,577 | 195.9 | 0.547 | 5,803,178 | 121.0 | 0.553 | 5,821,152 | 224.0 | 0.592 | 5,796,156 | 163.4 |
| **Age** | **1.100** |  |  | **1.093** |  |  | **1.104** |  |  | **1.098** |  |  |
| **SES quartile (ref. Disadvantaged)** |  | **2,105,793** | **484.9** |  | **2,161,269** | **299.1** |  | **2,095,583** | **555.8** |  | **2,154,805** | **398.1** |
| Int. low | 0.870 | 2,391,690 | 233.2 | 0.865 | 2,425,540 | 148.3 | 0.888 | 2,386,113 | 290.2 | 0.818 | 2,421,942 | 192.9 |
| Int. high | 0.777 | 2,548,680 | 152.2 | 0.724 | 2,574,867 | 91.9 | 0.739 | 2,544,800 | 175.1 | 0.739 | 2,572,501 | 128.0 |
| Advantaged | 0.599 | 2,462,251 | 69.2 | 0.590 | 2,475,318 | 45.4 | 0.563 | 2,460,547 | 82.9 | 0.567 | 2,474,195 | 60.9 |
| Unknown | 0.989 | 107,021 | 342.9 | 0.962 | 108,345 | 192.0 | 1.072 | 106,654 | 339.4 | 0.931 | 108,137 | 210.8 |
| Missing | 0.795 | 1,877,206 | 15.7 | 0.835 | 1,686,067 | 10.8 | 0.849 | 1,876,911 | 18.7 | 0.799 | 1,685,885 | 12.8 |
| **Household composition (ref. Couples with children)** |  | **5,503,935** | **26.5** |  | **5,508,626** | **19.0** |  | **5,377,863** | **37.8** |  | **5,387,650** | **27.1** |
| Isolated | 1.477 | 1,744,620 | 355.9 | 1.524 | 1,718,738 | 242.6 | 1.176 | 1,798,045 | 442.1 | 1.315 | 1,764,714 | 335.0 |
| Couples without children | 1.112 | 2,559,680 | 243.7 | 1.198 | 2,538,616 | 179.2 | 0.971 | 2,592,041 | 319.7 | 1.051 | 2,579,179 | 237.7 |
| Single-parent households | 1.557 | 1,317,053 | 74.6 | 1.764 | 1,307,845 | 57.0 | 1.266 | 1,321,359 | 94.2 | 1.500 | 1,306,669 | 77.1 |
| Others | 1.679 | 230,430 | 198.3 | 1.820 | 220,880 | 147.1 | 1.160 | 231,527 | 215.1 | 1.460 | 223,545 | 188.3 |
| Collectives | 6.034 | 136,923 | 4,884.5 | 4.185 | 136,701 | 2,272.8 | 2.725 | 149,773 | 3,849.8 | 2.392 | 155,708 | 2287.0 |
| **Nationality (ref. Belgians)** |  | **10,065,990** | **203.9** |  | **10,039,981** | **130.1** |  | **10,078,773** | **238.8** |  | **10,066,532** | **172.3** |
| French | 1.048 | 170,324 | 152.1 | 0.988 | 167,508 | 90.7 | 1.026 | 169,500 | 184.7 | 0.884 | 166,682 | 105.6 |
| Italians | 0.994 | 155,696 | 328.2 | 0.915 | 155,866 | 177.1 | 0.927 | 154,152 | 371.1 | 0.979 | 154,220 | 260.0 |
| Other Europeans | 0.887 | 669,172 | 76.7 | 0.871 | 649,429 | 50.2 | 0.794 | 659,945 | 81.8 | 0.809 | 637,559 | 63.4 |
| Africans | 0.945 | 191,037 | 64.9 | 0.844 | 188,137 | 37.2 | 0.951 | 180,414 | 85.9 | 0.794 | 175,380 | 51.9 |
| Asians | 1.061 | 177,072 | 46.3 | 0.863 | 170,389 | 27.0 | 1.055 | 169,159 | 62.1 | 0.873 | 162,161 | 38.9 |
| Other nationalities | 0.688 | 63,350 | 26.8 | 0.833 | 60,096 | 21.6 | 0.952 | 58,665 | 44.3 | 0.734 | 54,931 | 27.3 |
| Excess mortality at 20 km | 2.531 |  |  |  |  |  | 3.578 |  |  |  |  |  |
| Proportion of isolated people | 1.016 |  |  | 0.999 |  |  | 1.001 |  |  | 1.005 |  |  |
| Distance to hospital | 1.001 |  |  | 1.000 |  |  | 1.004 |  |  | 1.002 |  |  |
| Community density | 1.000 |  |  | 1.000 |  |  | 1.000 |  |  | 1.000 |  |  |
| Proportion of disadvantaged | 1.012 |  |  | 1.011 |  |  | 1.009 |  |  | 1.008 |  |  |
| **Urban hierarchy (ref. Conurbation)** |  | **1,539,081** | **189.3** |  | **1,526,309** | **94.7** |  | **1,521,928** | **176.9** |  | **1,511,842** | **129.3** |
| Suburb | 0.954 | 1,907,456 | 190.6 | 1.055 | 1,899,616 | 126.8 | 1.056 | 1,895,448 | 238.7 | 0.987 | 1,889,904 | 165.2 |
| ZMA | 0.878 | 1,658,605 | 211.9 | 0.989 | 1,650,103 | 128.2 | 1.047 | 1,653,922 | 235.3 | 0.970 | 1,647,242 | 174.8 |
| Small town | 0.903 | 2,708,417 | 204.0 | 1.029 | 2,694,209 | 130.4 | 1.043 | 2,710,111 | 240.0 | 0.976 | 2,698,282 | 172.8 |
| Rural (or other) | 0.891 | 3,679,082 | 175.1 | 1.046 | 3,661,169 | 121.8 | 1.050 | 3,689,199 | 221.3 | 0.972 | 3,670,195 | 160.0 |
| AIC | 235,269.7 |  |  | 169,539.1 |  |  | 277,578.9 |  |  | 216,392.5 |  |  |
| BIC | 235,683.1 |  |  | 169,938.2 |  |  | 277,992.3 |  |  | 216,791.5 |  |  |

**Table 2.**
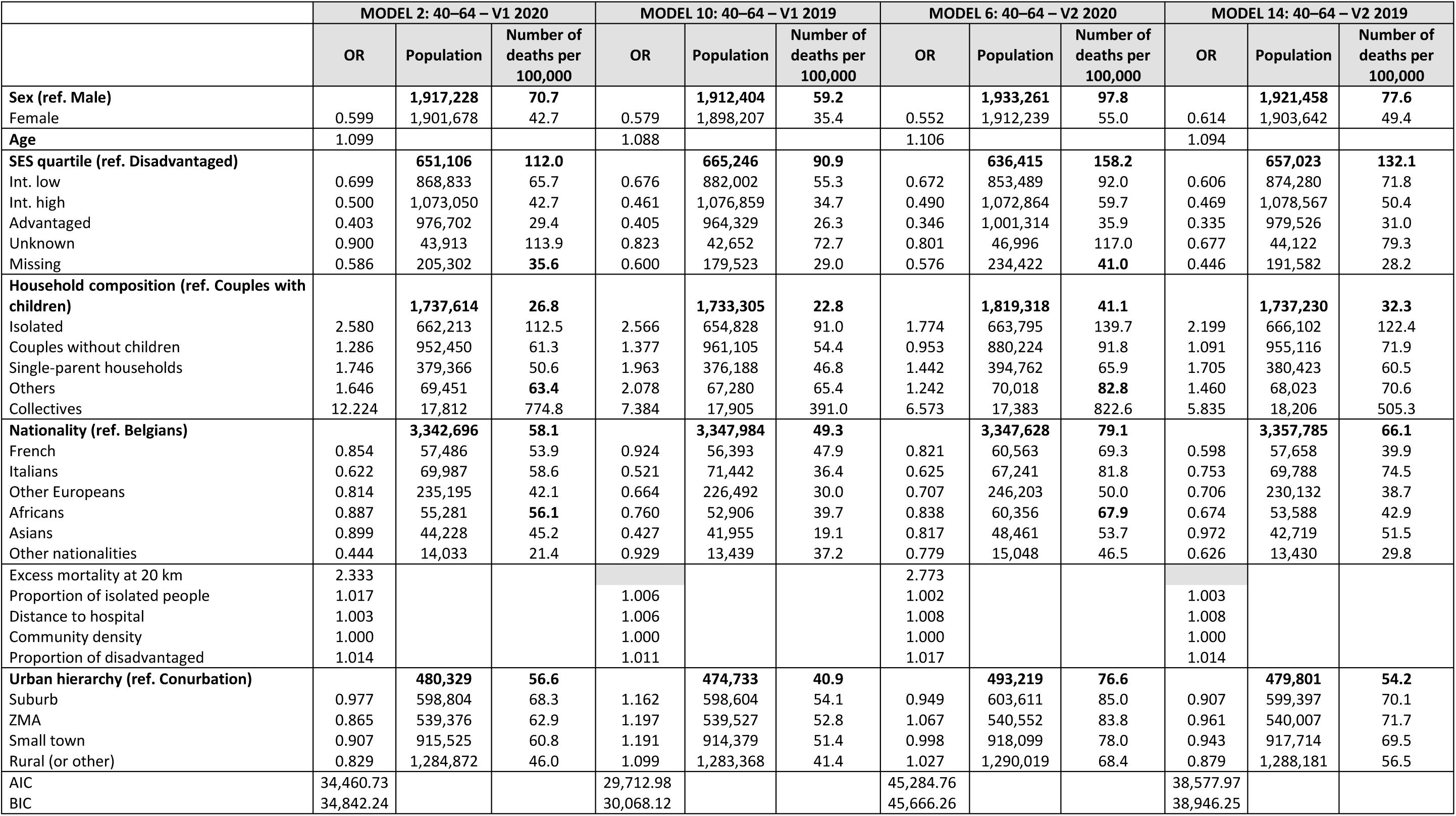
Multivariate results estimating the individual probability of dying (expressed in odds ratios) during one of the two Covid-19 waves (2020), compared with the results obtained during the same periods in 2019, models for people aged 40 to 64. Source: Statbel; authors’ calculations.

|  | MODEL 2: 40–64 – V1 2020 |  |  | MODEL 10: 40–64 – V1 2019 |  |  | MODEL 6: 40–64 – V2 2020 |  |  | MODEL 14: 40–64 – V2 2019 |  |  |
| --- | --- | --- | --- | --- | --- | --- | --- | --- | --- | --- | --- | --- |
|  | OR | Population | Number of deaths per 100,000 | OR | Population | Number of deaths per 100,000 | OR | Population | Number of deaths per 100,000 | OR | Population | Number of deaths per 100,000 |
| **Sex (ref. Male)** |  | **1,917,228** | **70.7** |  | **1,912,404** | **59.2** |  | **1,933,261** | **97.8** |  | **1,921,458** | **77.6** |
| Female | 0.599 | 1,901,678 | 42.7 | 0.579 | 1,898,207 | 35.4 | 0.552 | 1,912,239 | 55.0 | 0.614 | 1,903,642 | 49.4 |
| **Age** | **1.099** |  |  | **1.088** |  |  | **1.106** |  |  | **1.094** |  |  |
| **SES quartile (ref. Disadvantaged)** |  | **651,106** | **112.0** |  | **665,246** | **90.9** |  | **636,415** | **158.2** |  | **657,023** | **132.1** |
| Int. low | 0.699 | 868,833 | 65.7 | 0.676 | 882,002 | 55.3 | 0.672 | 853,489 | 92.0 | 0.606 | 874,280 | 71.8 |
| Int. high | 0.500 | 1,073,050 | 42.7 | 0.461 | 1,076,859 | 34.7 | 0.490 | 1,072,864 | 59.7 | 0.469 | 1,078,567 | 50.4 |
| Advantaged | 0.403 | 976,702 | 29.4 | 0.405 | 964,329 | 26.3 | 0.346 | 1,001,314 | 35.9 | 0.335 | 979,526 | 31.0 |
| Unknown | 0.900 | 43,913 | 113.9 | 0.823 | 42,652 | 72.7 | 0.801 | 46,996 | 117.0 | 0.677 | 44,122 | 79.3 |
| Missing | 0.586 | 205,302 | **35.6** | 0.600 | 179,523 | 29.0 | 0.576 | 234,422 | **41.0** | 0.446 | 191,582 | 28.2 |
| **Household composition (ref. Couples with children)** |  | **1,737,614** | **26.8** |  | **1,733,305** | **22.8** |  | **1,819,318** | **41.1** |  | **1,737,230** | **32.3** |
| Isolated | 2.580 | 662,213 | 112.5 | 2.566 | 654,828 | 91.0 | 1.774 | 663,795 | 139.7 | 2.199 | 666,102 | 122.4 |
| Couples without children | 1.286 | 952,450 | 61.3 | 1.377 | 961,105 | 54.4 | 0.953 | 880,224 | 91.8 | 1.091 | 955,116 | 71.9 |
| Single-parent households | 1.746 | 379,366 | 50.6 | 1.963 | 376,188 | 46.8 | 1.442 | 394,762 | 65.9 | 1.705 | 380,423 | 60.5 |
| Others | 1.646 | 69,451 | **63.4** | 2.078 | 67,280 | 65.4 | 1.242 | 70,018 | **82.8** | 1.460 | 68,023 | 70.6 |
| Collectives | 12.224 | 17,812 | 774.8 | 7.384 | 17,905 | 391.0 | 6.573 | 17,383 | 822.6 | 5.835 | 18,206 | 505.3 |
| **Nationality (ref. Belgians)** |  | **3,342,696** | **58.1** |  | **3,347,984** | **49.3** |  | **3,347,628** | **79.1** |  | **3,357,785** | **66.1** |
| French | 0.854 | 57,486 | 53.9 | 0.924 | 56,393 | 47.9 | 0.821 | 60,563 | 69.3 | 0.598 | 57,658 | 39.9 |
| Italians | 0.622 | 69,987 | 58.6 | 0.521 | 71,442 | 36.4 | 0.625 | 67,241 | 81.8 | 0.753 | 69,788 | 74.5 |
| Other Europeans | 0.814 | 235,195 | 42.1 | 0.664 | 226,492 | 30.0 | 0.707 | 246,203 | 50.0 | 0.706 | 230,132 | 38.7 |
| Africans | 0.887 | 55,281 | **56.1** | 0.760 | 52,906 | 39.7 | 0.838 | 60,356 | **67.9** | 0.674 | 53,588 | 42.9 |
| Asians | 0.899 | 44,228 | 45.2 | 0.427 | 41,955 | 19.1 | 0.817 | 48,461 | 53.7 | 0.972 | 42,719 | 51.5 |
| Other nationalities | 0.444 | 14,033 | 21.4 | 0.929 | 13,439 | 37.2 | 0.779 | 15,048 | 46.5 | 0.626 | 13,430 | 29.8 |
| Excess mortality at 20 km | 2.333 |  |  |  |  |  | 2.773 |  |  |  |  |  |
| Proportion of isolated people | 1.017 |  |  | 1.006 |  |  | 1.002 |  |  | 1.003 |  |  |
| Distance to hospital | 1.003 |  |  | 1.006 |  |  | 1.008 |  |  | 1.008 |  |  |
| Community density | 1.000 |  |  | 1.000 |  |  | 1.000 |  |  | 1.000 |  |  |
| Proportion of disadvantaged | 1.014 |  |  | 1.011 |  |  | 1.017 |  |  | 1.014 |  |  |
| **Urban hierarchy (ref. Conurbation)** |  | **480,329** | **56.6** |  | **474,733** | **40.9** |  | **493,219** | **76.6** |  | **479,801** | **54.2** |
| Suburb | 0.977 | 598,804 | 68.3 | 1.162 | 598,604 | 54.1 | 0.949 | 603,611 | 85.0 | 0.907 | 599,397 | 70.1 |
| ZMA | 0.865 | 539,376 | 62.9 | 1.197 | 539,527 | 52.8 | 1.067 | 540,552 | 83.8 | 0.961 | 540,007 | 71.7 |
| Small town | 0.907 | 915,525 | 60.8 | 1.191 | 914,379 | 51.4 | 0.998 | 918,099 | 78.0 | 0.943 | 917,714 | 69.5 |
| Rural (or other) | 0.829 | 1,284,872 | 46.0 | 1.099 | 1,283,368 | 41.4 | 1.027 | 1,290,019 | 68.4 | 0.879 | 1,288,181 | 56.5 |
| AIC | 34,460.73 |  |  | 29,712.98 |  |  | 45,284.76 |  |  | 38,577.97 |  |  |
| BIC | 34,842.24 |  |  | 30,068.12 |  |  | 45,666.26 |  |  | 38,946.25 |  |  |

**Table 3.** Multivariate results estimating the individual probability of dying (expressed in odds ratios) during one of the two Covid-19 waves (2020), compared with the results obtained during the same periods in 2019, models for people aged 65 to 79. Source: Statbel; authors’ calculations.

|  | MODEL 3: 65–79 – V1 2020 |  |  | MODEL 11: 65–79 – V1 2019 |  |  | MODEL 7: 65–79 – V2 2020 |  |  | MODEL 15: 65–79 – V2 2019 |  |  |
| --- | --- | --- | --- | --- | --- | --- | --- | --- | --- | --- | --- | --- |
|  | OR | Population | Number of deaths per 100,000 | OR | Population | Number of deaths per 100,000 | OR | Population | Number of deaths per 100,000 | OR | Population | Number of deaths per 100,000 |
| **Sex (ref. Male)** |  | **729,352** | **465.6** |  | **713,718** | **319.3** |  | **804,468** | **790.6** |  | **745,450** | **410.4** |
| Female | 0.507 | 818,339 | 281.9 | 0.503 | 804,594 | 188.8 | 0.502 | 884,093 | 1,027.0 | 0.523 | 829,384 | 252.7 |
| **Age** | 1.099 |  |  | 1.080 |  |  | 1.100 |  |  | 1.088 |  |  |
| **SES quartile (ref. Disadvantaged)** |  | **449,161** | **527.0** |  | **461,983** | **334.6** |  | **455,559** | **628.9** |  | **460,592** | **471.8** |
| Int. low | 0.828 | 457,358 | 344.2 | 0.851 | 450,784 | 239.1 | 0.833 | 492,049 | 416.0 | 0.784 | 465,191 | 309.8 |
| Int. high | 0.698 | 358,016 | 288.0 | 0.716 | 339,642 | 203.2 | 0.698 | 412,986 | 318.4 | 0.661 | 363,511 | 252.3 |
| Advantaged | 0.536 | 254,008 | 212.6 | 0.607 | 239,240 | 166.8 | 0.502 | 294,234 | 225.7 | 0.532 | 256,959 | 199.3 |
| Unknown | 0.910 | 10,379 | 1,156.2 | 0.773 | 10,373 | 510.9 | 1.095 | 11,316 | 1,069.3 | 0.849 | 10,786 | 602.6 |
| Missing | 0.759 | 18,769 | 378.3 | 0.620 | 16,290 | 196.4 | 0.864 | 22,417 | 432.7 | 0.684 | 17,795 | 264.1 |
| **Household composition (ref. Couples with children)** |  | **133,805** | **325.1** |  | **131,119** | **193.7** |  | **155,393** | **400.3** |  | **135,792** | **284.3** |
| Isolated | 1.249 | 397,344 | 394.6 | 1.526 | 387,555 | 280.0 | 0.985 | 431,540 | 452.8 | 1.214 | 405,289 | 366.2 |
| Couples without children | 0.848 | 916,546 | 265.8 | 1.054 | 901,190 | 201.6 | 0.783 | 991,376 | 344.1 | 0.912 | 929,813 | 272.1 |
| Single-parent households | 1.287 | 50,693 | 382.7 | 1.642 | 49,790 | 283.2 | 1.054 | 57,185 | 426.7 | 1.253 | 51,687 | 346.3 |
| Others | 1.256 | 25,625 | 405.9 | 1.773 | 24,994 | 332.1 | 0.959 | 28,267 | 431.6 | 1.278 | 26,111 | 383.0 |
| Collectives | 10.312 | 23,678 | 4,079.7 | 7.823 | 23,664 | 1,766.4 | 4.428 | 24,800 | 3,048.4 | 4.306 | 26,142 | 1,820.8 |
| **Nationality (ref. Belgians)** |  | **1,442,626** | **366.7** |  | **1,414,889** | **250.1** |  | **1,572,033** | **417.4** |  | **1,467,569** | **325.4** |
| French | 0.797 | 16,656 | 372.2 | 0.968 | 16,421 | 274.0 | 1.081 | 18,363 | 582.7 | 1.141 | 17,084 | 415.6 |
| Italians | 0.891 | 31,259 | 451.1 | 0.825 | 30,725 | 257.1 | 0.899 | 34,103 | 539.5 | 1.083 | 31,879 | 448.6 |
| Other Europeans | 0.877 | 42,491 | 346.0 | 0.951 | 41,666 | 242.4 | 0.745 | 47,922 | 321.4 | 0.840 | 43,499 | 280.5 |
| Africans | 0.884 | 7,566 | 462.6 | 0.759 | 7,645 | 222.4 | 1.110 | 8,001 | 674.9 | 0.727 | 7,646 | 300.8 |
| Asians | 1.045 | 5,584 | 411.9 | 1.165 | 5,482 | 291.9 | 1.360 | 6,395 | 625.5 | 0.867 | 5,616 | 302.7 |
| Other nationalities | 0.664 | 1,509 | 331.3 | 0.528 | 1,484 | 134.8 | 1.000 | 1,744 | 458.7 | 0.585 | 1,541 | 194.7 |
| Excess mortality at 20 km | 2.647 |  |  |  |  |  | 3.383 |  |  |  |  |  |
| Proportion of isolated people | 1.020 |  |  | 0.992 |  |  | 1.007 |  |  | 1.004 |  |  |
| Distance to hospital | 1.003 |  |  | 0.996 |  |  | 1.007 |  |  | 1.007 |  |  |
| Community density | 1.000 |  |  | 1.000 |  |  | 1.000 |  |  | 1.000 |  |  |
| Proportion of disadvantaged | 1.011 |  |  | 1.014 |  |  | 1.013 |  |  | 1.009 |  |  |
| **Urban hierarchy (ref. Conurbation)** |  | **150,733** | **500.2** |  | **149,127** | **274.9** |  | **163,862** | **431.5** |  | **153,870** | **338.6** |
| Suburb | 0.876 | 242,365 | 385.0 | 0.992 | 238,860 | 278.4 | 1.176 | 263,017 | 489.7 | 0.994 | 246,979 | 373.7 |
| ZMA | 0.889 | 232,685 | 390.7 | 0.855 | 228,340 | 236.5 | 1.158 | 253,452 | 434.0 | 0.930 | 236,705 | 330.8 |
| Small town | 0.898 | 392,236 | 375.3 | 0.942 | 384,331 | 249.8 | 1.147 | 427,822 | 427.3 | 0.918 | 399,176 | 328.4 |
| Rural (or other) | 0.843 | 529,672 | 308.7 | 0.966 | 517,654 | 236.3 | 1.122 | 580,408 | 376.6 | 0.884 | 538,104 | 300.5 |
| AIC | 70,200.6 |  |  | 51,150.4 |  |  | 86,629.3 |  |  | 66,644.5 |  |  |
| BIC | 70,555.9 |  |  | 51,493.0 |  |  | 86,984.5 |  |  | 66,987.0 |  |  |

**Table 4.** Multivariate results estimating the individual probability of dying (expressed in odds ratios) during one of the two Covid-19 waves (2020), compared with the results obtained during the same periods in 2019, models for people aged 80 and over. Source: Statbel; authors’ calculations.

|  | MODEL 4: 80+ – V1 2020 |  |  | MODEL 12: 80+ – V1 2019 |  |  | MODEL 8: 80+ – V2 2020 |  |  | MODEL 16: 80+ – V2 2019 |  |  |
| --- | --- | --- | --- | --- | --- | --- | --- | --- | --- | --- | --- | --- |
|  | OR | Population | Number of deaths per 100,000 | OR | Population | Number of deaths per 100,000 | OR | Population | Number of deaths per 100,000 | OR | Population | Number of deaths per 100,000 |
| **Sex (ref. Male)** |  | **238,795** | **2,386.1** |  | **235,465** | **1,414.2** |  | **265,965** | **2,391.3** |  | **261,690** | **1,633.6** |
| Female | 0.570 | 408,193 | 2,014.2 | 0.616 | 403,443 | 1,177.4 | 0.637 | 444,913 | 2,040.8 | 0.694 | 438,568 | 1,446.8 |
| **Age** | 1.090 |  |  | 1.108 |  |  | 1.101 |  |  | 1.108 |  |  |
| **SES quartile (ref. Disadvantaged)** |  | **299,155** | **2,358.3** |  | **297,740** | **1,425.1** |  | **342,182** | **2,243.8** |  | **321,679** | **1,698.6** |
| Int. low | 0.932 | 176,168 | 1,911.8 | 0.937 | 168,748 | 1,168.6 | 0.984 | 208,652 | 1,929.5 | 0.896 | 186,809 | 1,360.7 |
| Int. jigh | 0.925 | 115,595 | 2,026.9 | 0.852 | 111,768 | 1,128.2 | 0.869 | 136,167 | 1,794.1 | 0.898 | 122,764 | 1,440.2 |
| Advantaged | 0.786 | 56,412 | 1,519.2 | 0.710 | 51,416 | 846.0 | 0.779 | 71,310 | 1,377.1 | 0.773 | 59,046 | 1,102.5 |
| Unknown | 0.874 | 5,347 | 3,572.1 | 0.959 | 5,671 | 2,098.4 | 0.974 | 5,997 | 3,034.9 | 0.904 | 6,037 | 2,037.4 |
| Missing | 0.893 | 4,110 | 2,579.1 | 0.917 | 3,565 | 1,402.5 | 1.136 | 5,014 | 2,572.8 | 1.151 | 3,923 | 1,835.3 |
| **Household composition (ref. Couples with children)** |  | **26,996** | **1,622.5** |  | **26,962** | **1,031.1** |  | **31,968** | **1,701.7** |  | **28,590** | **1,360.6** |
| Isolated | 0.939 | 261,338 | 1,471.7 | 0.892 | 255,515 | 952.6 | 0.858 | 300,685 | 1,663.2 | 0.842 | 274,944 | 1,291.2 |
| Couples without children | 0.862 | 239,618 | 1,333.8 | 0.937 | 231,354 | 944.0 | 0.859 | 287,353 | 1,405.2 | 0.878 | 251,033 | 1,150.4 |
| Single-parent households | 1.128 | 31,091 | 1,797.9 | 1.144 | 30,826 | 1,239.2 | 0.958 | 35,727 | 1,914.5 | 1.039 | 33,141 | 1,629.4 |
| Others | 1.292 | 14,165 | 2,132.0 | 1.173 | 14,149 | 1,321.6 | 0.930 | 16,152 | 1,931.6 | 1.062 | 15,196 | 1,730.7 |
| Collectives | 3.710 | 83,579 | 6,675.1 | 2.313 | 80,102 | 3,264.6 | 1.876 | 97,437 | 4,988.9 | 1.434 | 97,354 | 3,071.3 |
| **Nationality (ref. Belgians)** |  | **621,478** | **2,105.9** |  | **605,038** | **1,265.0** |  | **726,349** | **2,010.5** |  | **662,669** | **1,523.8** |
| French | 1.272 | 5,224 | 3,101.1 | 1.030 | 4,973 | 1,508.1 | 1.051 | 6,071 | 2,684.9 | 0.804 | 5,473 | 1,425.2 |
| Italians | 1.192 | 11,942 | 2,755.0 | 1.164 | 11,520 | 1,484.4 | 1.103 | 14,392 | 2,306.8 | 1.039 | 12,771 | 1,589.5 |
| Other Europeans | 0.957 | 12,896 | 1,961.8 | 0.932 | 12,295 | 1,098.0 | 0.860 | 16,048 | 1,545.4 | 0.899 | 13,727 | 1,311.3 |
| Africans | 0.832 | 3,213 | 1,556.2 | 0.824 | 3,116 | 866.5 | 0.724 | 3,978 | 1,282.1 | 0.731 | 3,429 | 1,049.9 |
| Asians | 1.079 | 1,669 | 1,917.3 | 0.888 | 1,603 | 935.7 | 0.957 | 2,055 | 1,654.5 | 0.786 | 1,800 | 1,111.1 |
| Other nationalities | 0.656 | 365 | 1,643.8 | 0.614 | 363 | 826.4 | 0.960 | 429 | 2,097.9 | 0.718 | 389 | 1,285.3 |
| Excess mortality at 20 km | 2.546 |  |  |  |  |  | 3.860 |  |  |  |  |  |
| Proportion of isolated people | 1.008 |  |  | 0.996 |  |  | 0.992 |  |  | 1.002 |  |  |
| Distance to hospital | 0.997 |  |  | 1.000 |  |  | 1.001 |  |  | 0.998 |  |  |
| Community density | 1.000 |  |  | 1.000 |  |  | 1.000 |  |  | 1.000 |  |  |
| Proportion of disadvantaged | 1.011 |  |  | 1.009 |  |  | 1.006 |  |  | 1.004 |  |  |
| **Urban hierarchy (ref. Conurbation)** |  | **68,399** | **2,704.7** |  | **67,590** | **1,195.4** |  | **78,888** | **1,990.2** |  | **73,189** | **1,557.6** |
| Suburb | 1.001 | 107,313 | 2,092.9 | 1.057 | 105,337 | 1,304.4 | 1.078 | 124,926 | 2,130.1 | 1.027 | 114,799 | 1,507.0 |
| ZMA | 0.885 | 102,236 | 2,181.2 | 1.023 | 99,698 | 1,243.8 | 1.050 | 119,193 | 1,921.3 | 1.034 | 108,919 | 1,525.9 |
| Small town | 0.930 | 165,523 | 2,084.9 | 1.072 | 160,191 | 1,266.6 | 1.071 | 194,082 | 2,010.0 | 1.069 | 176,133 | 1,500.0 |
| Rural (or other) | 0.942 | 213,316 | 1,942.2 | 1.103 | 206,092 | 1,275.6 | 1.101 | 252,233 | 1,989.4 | 1.106 | 227,218 | 1,516.6 |
| AIC | 124,432.3 |  |  | 82,695.1 |  |  | 138,800.3 |  |  | 104,439.5 |  |  |
| BIC | 124,762.8 |  |  | 83,013.7 |  |  | 139,130.1 |  |  | 104,757.8 |  |  |

At the same time, a one-year increase in age leads to an additional risk of death of around 10%. But here again, this is not a characteristic of the pandemic period: the frailty of the elderly appears to be equivalent in 2019 and 2020.

In addition, a social gradient is observed: the more socially disadvantaged individuals are, the greater their risk of death. In 2020, compared with the most disadvantaged social group, the privileged class had an odds ratio at least 40% lower (in wave 1, OR = 0.599; in wave 2, OR = 0.563). However, the social gradient observed does not differ from that observed in conventional mortality either, where the most disadvantaged social groups are those which, all other things being equal, are characterised by a higher odd of death. In other words, the pandemic period neither partially erased nor significantly increased social differences in mortality, with the result that the social effect of Covid-19-related mortality was not neutral.

Note the case of people for whom the social group cannot be determined (unknown social group): in terms of mortality, they behave more or less identically to the most disadvantaged social group, in times of pandemic or not. This ‘all other things being equal’ finding is interesting insofar as, in the context of descriptive analyses, we had already mentioned the excess mortality of the ‘undetermined’ social group during the Covid waves of 2020 (Bourguignon et al., 2021). This can be explained in part by the fact that, from a socio-economic point of view, this group is also *a priori* in a precarious situation, characterised in particular by an overrepresentation of people of immigrant origin and people living in the Brussels Region (Bourguignon et al., 2021). However, overall, nationality does not lead to substantial differences in mortality.

Finally, the household composition of individuals generates differences in mortality. According to the regression models, individuals living in a couple with or without children would be the least exposed to mortality during the two Covid waves in 2020. On the other hand, the differences are more marked for other types of households. For example, the excess mortality of single individuals or single-parent households is confirmed in 2020, in the same way as that observed for usual mortality (2019). The differences between 2019 and 2020 relate more to collective households, for which mortality was amplified in 2020. In the first wave, residents of collective households had a risk of death around six times higher than that observed for couples with (a) child(ren) (OR in 2019 – equivalent to wave 1 – is around 4 times higher). In the second Covid wave, excess mortality still characterised these collective households but was roughly equivalent (OR = 2.7) to that observed in 2019 during the similar period (OR = 2.4).

Among the contextual variables, the intensity of excess local mortality affects the individual odd of dying in 2020. During the two waves of Covid-19, a one-unit increase in the local excess mortality indicator – i.e., a massive increase in mortality, which is not observed in reality^11^ – is associated with an individual risk of dying 2.5 times and 3.5 times higher in the first and second waves respectively. At the same time, a 1 percentage point increase in the proportions of isolated or disadvantaged people results in a 1% increase (OR = 1.01) in the individual risk of death. The effect of these variables may seem smaller than that of excess local mortality. Still, the amplitude of the values observed for the variables can have a significant effect on the individual risk of dying^12^. The other contextual variables (population density, distance from the nearest hospital) result in minor differences in mortality, both in a pandemic context and in a ‘classic’ mortality context.

### Characteristics related to mortality by large age group

The results distinguished by large age group remain approximately the same, as the individual characteristics associated with mortality remain very close from one group to the other. Estimates suggest that the probability of dying during the Covid-19 period is related to factors similar to those observed in usual mortality (2019). Whatever the age group, therefore, gender, age, and social affiliation generate similar mortality differentials, whether during a pandemic or not. There are, however, several specificities for certain age groups that merit highlighting.

Firstly, the effect of belonging to a collective household may vary between different age groups. The risk of mortality remains higher for individuals living in collective households, but more so for those aged 65–79 (OR = 10.3 (wave 1) or 4.4 (wave 2)) than for those aged 80 and over (OR = 3.7 (wave 1) or 1.9 (wave 2))^13^. This situation according to age (65–79 vs. 80+) and according to period (wave 1 vs. wave 2) is probably explained by a double selection effect: on the one hand, those who survive the first wave would be more robust than those who have died; on the other hand, those who survive the very old ages (80 and over) would also be potentially selected, because of a better state of health which would have enabled them to survive to that point. Nevertheless, such a difference in coefficients between two logistic models can be explained by the unobserved heterogeneity of mortality between different age groups (Mood, 2010).

Secondly, the effect of excess local mortality increases among the oldest populations. This is particularly true for the second wave of Covid-19, with the effect of excess local mortality on the individual odd of dying increasing with age. This could be partly explained by the fact that, even if the transmission mechanisms of the virus that generated the pandemic were a little better understood during the second wave, the relaxation of some health measures (or of compliance with them) could have led to significant outbreaks of excess mortality, with fatal consequences for the oldest age groups.

## Discussion

The main objectives of this article are (1) to measure the relative importance of individual and spatial determinants of mortality during and outside the Covid-19 pandemic, and (2) to measure differences in mortality according to individual and spatial characteristics. The combination of these two approaches has provided an original viewpoint for analysing the determinants of mortality during a pandemic while offering a comparison of mortality patterns over time (2019 vs. 2020), according to the evolution of the pandemic (wave 1 vs. wave 2) and according to the age of individuals (40–64 years, 65–79 years and 80 years and over).

The data for this purpose come from the National Register, the 2017 tax register, and the 2011 census. There are many advantages to using these data. In particular, the exhaustive nature of these sources allows an analysis of what mortality was like in 2019 and 2020, without the bias associated with inferring and generalising results from population samples.

As a reminder, three research hypotheses were mooted based on abundant international literature. The results obtained allow us to respond to them and qualify them based on the case of Belgium.

### First hypothesis: The relative weight of the variables

Firstly, we postulated that the relative importance of individual and spatial determinants of mortality would be modified in a pandemic context, with an increase in the weight of contextual variables and a change in the hierarchy of predominant variables, given the highly contagious nature of the virus associated with the Covid-19 pandemic. At this stage, the hypothesis has not been confirmed: the probability of dying during the Covid-in 2020 period is mainly influenced by the same individual variables as during similar periods in 2019, i.e., age, sex, and household composition. Nevertheless, some spatial variables played a decisive role in mortality in 2020, such as the local excess mortality index and the proportions of disadvantaged or isolated people in the municipality of residence. The extent of excess local mortality is in line with what has been observed in France (Levratto et al., 2020) and China (Kang et al., 2021). On the other hand, the results obtained contradict those of other studies that show a greater vulnerability during a pandemic period of individuals living in densely populated regions of France (Pilkington et al., 2021) and in the United States (Abedi et al., 2021).

The other individual variables (nationality, social group) and spatial variables (population density, distance to the nearest hospital) have a minimal influence on mortality, even in a pandemic. Compared with 2019, there is no real change in the hierarchy of variables according to their respective weight on mortality. Furthermore, the weight of contextual variables does not override that of individual variables, whether in the Covid-19 period or not.

### Second hypothesis: The characteristics associated with mortality

The second hypothesis involved identifying mortality-related characteristics during the Covid-19 pandemic. The vulnerability of some population groups that were already fragile in a context of traditional mortality would have been amplified during the pandemic, for example, men, older people, collective households, and disadvantaged social groups.

Overall, and with a few exceptions, our hypothesis has not been confirmed. Age and sex relate to multiple, well-studied phenomena, combining biology, metabolic state, health behaviours, and lifestyles. The year 2020 is no exception to the rule of a significant increase in the risk of death after the first year of life, as it is no exception to the excess male mortality rate up to the oldest ages. Social inequalities in mortality neither increased nor decreased during the Covid-19 crisis, which means that the social effect of Covid-19-related mortality is not neutral. It is apparent at the individual level in the excess mortality of the disadvantaged social group and in the existence of a social gradient in mortality. It is also apparent at the municipal level, with a non-negligible effect from variables such as the proportions of disadvantaged people. The pandemic would thus have perpetuated existing and significant disparities to the detriment of disadvantaged populations, as observed elsewhere in the world (Barhoumi et al., 2020; Chen & Krieger, 2021; Mayer, 2022).

However, there is one point on which the hypothesis is confirmed: the vulnerability of collective households during a pandemic. The excess mortality in collective households also reflects, to some extent, the determining effect of biological factors on mortality via age and/or state of health. It is also accepted that other factors may have contributed to the ‘vulnerable state’ characteristic of residents of collective households (proximity between residents and with carers, a particularly contagious virus (Davidson & Szanton, 2020; Kemenesi et al., 2020), and the lack of resources, particularly masks and hydroalcoholic gels).

### Third hypothesis: The specific characteristics of the large age groups

Finally, the third hypothesis addressed possible differences in mortality patterns by large age groups before and during the Covid-19 pandemic. Overall, this hypothesis was not confirmed, with relatively limited differences between the major age groups, both in terms of the weight of the variables and the disparities between population subgroups. With a few nuances between age groups, age and sex are consistently among the main determinants of mortality patterns, whatever the age group; in other words, the classic biological factors of age and sex act identically across the age groups.

In particular, the hypothesis predicted that the social background of individuals would have a more visible impact on mortality before the age of 65, given that mortality before this age is considered ‘avoidable’ or ‘premature’, with, *a priori*, the biological effects of age and sex being less significant than at older ages. However, even during a pandemic, there was no reduction or increase in the mortality gap.

At older ages, on the other hand, we hypothesised that a biological effect would supplant the social effect, with a return of age, sex, and state of health (via household composition) as determining variables. In reality, the only notable difference that emerges is in the variable of household composition. Its weight increases steadily with the age of individuals until it becomes the variable with the highest weight at age 65 and over in the first Covid wave of 2020. Analyses of the parameters for each modality show a remarkable risk of death for residents of collective households, and more so for those aged 65–79 than for those aged 80 and over. This is probably a selection effect. According to Pujol et al. (2021), compared with the population living in private households, the risk of death in a nursing home is much higher among relatively young residents (65–74) than among those aged 75 and over. For people aged 65–79, institutionalisation likely results from health problems that prevent them from remaining at home^14^. Among those aged 80 and over, nursing home residents have survived to a ripe old age and, in addition to medical care, for some of them, residence in an institution is a response not only to health issues but to a need for social interaction and contact (Hanratty et al., 2018), even if this need is not systematically met (Gardiner et al., 2020). The age, sex, and household situation of individuals would thus have been major determinants of mortality during the Covid period of 2020. Alongside these variables, living close to areas of excess mortality would determine the risk of death, particularly at older ages. As this is an indicator of the virulence of the virus in the area, this implies that the individual risk of dying during the Covid period is increased in elderly people living in an area of high mortality, thus pointing to high circulation of the virus. The spatial context of mortality would therefore have repercussions on the individual risk of mortality, and this is all the more justified in the case of the elderly as their resistance and immune capacity diminish. However, comparing the coefficients of logistic regressions carried out on different population groups is not easy, since the difference in coefficients can be explained by unobservable phenomena, which have different impacts on mortality depending on the age group (Mood, 2010).

## General conclusion

In general, and in conclusion, the results show that, with a few nuances, non-pandemic mortality patterns do not differ from those observed during the Covid pandemic. This can be explained by the fact that vulnerability to the Covid-19 virus mainly affected population groups that were already the most vulnerable under normal circumstances (elderly people, men, nursing home residents). In addition to the almost identical mortality pattern, it is essential to emphasise that during a pandemic, there is an almost systematic increase in partial pseudo-correlations. The combination of these two factors allows us to emphasise that the pandemic amplified inequalities in mortality in certain groups, which, even before the pandemic, were already paying a heavy price in terms of mortality.

## Limitations and methodological aspects

Several limitations may affect our analyses over and above the results and the conclusions we can draw from them. Firstly, it should be remembered that the analyses relate to all-cause mortality during the two Covid waves in 2020. Currently, data by cause of death are unavailable in Belgium, making it impossible to identify deaths that resulted directly from the pandemic. If Covid-19 doubled the number of deaths during the first wave of 2020, a certain number of ‘all-cause’ deaths were due to causes other than Covid-19.

There are also inherent limitations in the indicators used. For example, we have tried to explain the probability of dying in 2019 or 2020 by characteristics observed much earlier. This is the case for the social positioning of individuals, which is based on data from 2011 and 2017. However, individual situations may have changed between 2011 or 2017 and the most recent period, even if this problem probably affects the oldest individuals much less. At older ages, for whom mortality was highest, education levels, income, and access to housing are more or less stable (Eggerickx et al., 2020). There is no alternative for addressing this concern, as the next update is expected to take place in 2021, i.e., at a later date than that relating to the events studied, and with no possible adjustment for people who died between 2011 and 2021.

The possibilities for research into Covid-19 are far from exhausted. The availability of data by cause of death will offer other possibilities for analysis, with questions that could be even more precise since we will be able to work solely with data on the victims of the pandemic. Still, analyses on all-cause excess mortality will remain solid on the consequences of the pandemic, in terms of death directly related to Covid, as well as other causes of death.

## Data Availability

No data produced in the present study are available

## Footnotes

1 In Belgium, both Covid-certified and -suspected deaths were considered as Covid deaths, whether they occurred in hospital, at home or in a nursing home.

2 The study used a multidimensional index incorporating four indicators: level of education, income, employment category, and housing characteristics. For more details, see the methodology section of this article.

3 This is the ratio between the age-standardised mortality rate calculated for the two 2020 waves and the standardised mortality rate calculated for the same months over the 2016–2019 observation period.

4 We chose the method of smoothing by potential, i.e., Stewart’s method (Grasland, Mathian, and Vincent 2000; Stewart and Warntz 1958), which we performed using the online interface Magrit (http://magrit.cnrs.fr). The following parameters were used: exponential function, span = 10 km, beta = 2.

5 MR(S) stands for ‘Maisons de repos et de soins’, the appellation for Belgian nursing and care homes.

6 The most precarious social groups more often carry out front-line jobs, and strict compliance with health measures depends on living conditions (e.g., high household density).

7 The first wave runs from 15 March to 30 April 2020 and the second wave from 15 October to 31 December 2020.

8 The distance between the commune and the nearest hospital is calculated from the centroid of the individual communes of residence. It therefore does not include information on travel time which, in specific rural contexts in the south of Belgium, for example, can also indicate other inequalities in healthcare access.

9 Urban hierarchy is based on the typology of urban residential complexes developed by Vanderstraeten and Van Hecke (2019). It distinguishes five types of place: the conurbation, suburbs (urbanised belt of around urban centre), ZMAs (*zones des migrants alternants*, areas outside cities with a substantial commuter population, small towns, and the rural environment (areas not classified within the urban hierarchy).

10 The under-40s were not considered, as no excess mortality in this population was observed in 2020 in the descriptive analyses (Bourguignon et al., 2022).

11 At the local level, the excess mortality indices do not exceed 1.7 in wave 1 and 1.4 in wave 2. A local excess mortality of 2 would mean that mortality in 2020 would have doubled compared with 2019.

12 The difference between the lowest and highest proportion of isolated people at the municipal level is 28 percentage points (minimum value = 7.79%; maximum value = 36.04%). Regarding the proportions of disadvantaged people, the difference between the minimum and maximum proportions is 56 percentage points (minimum value = 6.54%; maximum value = 62.87%), with potentially significant differences in the risk of death between individuals belonging to different municipalities.

13 We are not commenting here on the odds ratios for residents of collective households before the age of 65, who are inevitably less affected by factors related to old age and proportionally very few (on 1 Jan. 2020, 0.5% of 40– 64-year-olds were living in collective households, compared with 1.5% of 65–79-year-olds and 12.7% of those aged 80 and over).

14 In Brussels, the risk of death in a nursing home is strongly associated with age and pathologies, such as chronic illnesses (Houttekier et al., 2009).

